# Shatavari Supplementation in Postmenopausal Women Improves Handgrip Strength and Increases *Vastus Lateralis* Myosin Regulatory Light Chain Phosphorylation But Does Not Alter Markers of Bone Turnover: A Randomised Controlled Trial

**DOI:** 10.1101/2021.09.16.21263687

**Authors:** Mary F. O’Leary, Sarah R. Jackman, Vlad R. Sabou, Matthew I Campbell, Jonathan C. Y. Tang, John Dutton, Joanna L. Bowtell

**Affiliations:** College of Life and Environmental Sciences, Sport and Health Sciences, Exeter University, Exeter, United Kingdom; Norwich Medical School, University of East Anglia, Norwich Research Park, Norwich, Norfolk, NR4 7TJ, UK

**Keywords:** asparagus racemosus, skeletal muscle, nutrition

## Abstract

**Background:** Shatavari has long been used as an Ayurvedic herb for women’s health, but empirical evidence for its effectiveness has been lacking. Shatavari contains phytoestrogenic compounds that bind to the estradiol receptor, and may therefore benefit postmenopausal women since postmenopausal estradiol deficiency contributes to sarcopenia and osteoporosis.

**Methods:** In a randomised double-blind trial, 20 postmenopausal women (68.5 ± 6 y) ingested either placebo (N=10) or shatavari (N=10; 1000 mg/d, equivalent to 26,500 mg/d fresh weight shatavari) for 6 weeks. Handgrip and knee extensor strength were measured at baseline and at 6 weeks. Vastus lateralis (VL) biopsy samples were obtained. Data are presented and analysed (t test/Mann Whitney U) as difference scores (Week 6 – baseline, median ± interquartile range).

**Results:** Handgrip, (but not knee extensor) strength was improved by shatavari supplementation (shatavari +0.7 ± 1.1 kg, placebo -0.4 ± 1.3 kg; p=0.04). Myosin regulatory light chain phosphorylation, a known marker of improved myosin contractile function, was increased in VL following shatavari supplementation (immunoblotting; placebo -0.08 ± 0.5 a.u. shatavari +0.3 ± 1 arbitrary units (a.u.); p=0.03). Shatavari increased phosphorylation of Aktser473 (Aktser473 (placebo -0.6 ± 0.6 a.u. shatavari +0.2 ± 1.3 a.u; p=0.03) in VL. Shatavari supplementation did not alter plasma markers of bone turnover (P1NP, β-CTX) and stimulation of human osteoblasts with pooled sera (N=8 per condition) from placebo and shatavari supplementation conditions did not alter cytokine or metabolic markers of osteoblast activity.

**Conclusions:** Shatavari may improve muscle function and contractility via myosin conformational change and warrants further investigation of its utility in conserving musculoskeletal function in postmenopausal women.

**Trial Registration:** Retrospectively registered at clinicaltrials.gov as NCT05025917 on 30/08/21.

## Background

Shatavari (*Asparagus Racemosus Willd*.) has long been used as an Ayurvedic herb for women’s health [1], but empirical evidence for its effectiveness remains limited. Steroidal saponins are thought to be the principal bioactive constituents of shatavari root. These saponins are known as shatavarins I–IV, and they are glycosides of sarsasapogenin [2]. Shatavari root also contains other chemical constituents of note, including racemosides, racemosol, racemofuran and asparagamine A, all of which display antioxidant activity [2]. The limited research that exists suggests that shatavari also contains phytoestrogenic compounds that are capable of binding to the estradiol (E2) receptors (E2R). Indeed, E2R functionalised magnetic nanoparticles can isolate phytoestrogens of 4.6 nmol E2-equivalent activity from a 1-mg crude extract of *Asparagus racemosus* [3]. The phytoestrogens that are most relevant to human health are those that are 1) distinct from structures that occur naturally in mammals 2) contained in plants that are consumed by humans and 3) bioavailable and capable of binding to human E2R [4]. Thus, the isoflavones genistein and daidzein, the coumestan coumestrol, the lignans enterolactone and enterodiol, the flavonols quercetin, kaempferol, rutin and the stilbene resveratrol represent the most prolific and relevant dietary forms of phytoestrogens in humans [4,5]. *In silico* research has suggested that shatavari-derived phytoestrogens including rutin, kaempferol, genistein, daidzein and quercetin bind to E2R with greater affinity than a selective estrogen receptor modulator control, bazedoxifene [6].

Shatavari administration in rats induces pro-estrogenic changes in mammary glands and genital organs [7]. Shatavari also appears to have galactogogue activity; in a placebo-controlled study that administered 60 mg/kg/d shatavari root powder to breastfeeding mothers for 30 days, systemic prolactin concentrations were increased (3-fold) and shatavari promoted infant weight gain [8]. However, a similar study, (in which the shatavari dose is unclear) found no effect on systemic prolactin concentrations [9].; both studies were of poor quality. The mechanism by which shatavari increases serum prolactin concentration is unknown. Little is definitively known about how herbal galactagogues act, beyond speculation that they have E2 like activity and rely upon phytoestrogen interaction with Erα. [10,11].

The potential for estrogen-like effects of shatavari supplementation has implications for both muscle and bone health after the menopause. Observational data have long suggested that in women, post-menopausal estrogenic deficits may promote the age-associated decline in muscle strength and function (sarcopenia); men experience a more gradual decline in handgrip strength (HGS) across the lifespan, whereas women display a sharp decline in such strength in the middle of the sixth decade [12]. A large meta-analysis of studies that have examined muscle strength and hormone replacement therapy (HRT) demonstrated that HRT increases muscle strength [13]. E2 withdrawal has a detrimental effect on myosin binding function and therefore muscle force production; conversely E2 improves muscle force production, and may promote muscle protein synthesis [14–16]. One study has shown that shatavari (500 mg·d^−1^ root extract; fresh weight equivalent not stated) enhanced strength gains in young men following eight weeks of bench press training [17]; this was an observational study with no indication of possible mechanisms of action. However, it does suggest that shatavari may be a beneficial ergogenic aid in supporting resistance training-induced increases in muscle size and strength; no assessment of these ergogenic effects has been made in older adults.

The decreased E2 production that precipitates menopause is also associated with a decline in bone mineral density, a phenomenon that increases the risk of bone fractures [18]. Elderly individuals that experience a hip fracture have a 3-fold increase in their risk of all-cause mortality [19]. HRT decreases the risk of osteoporosis and fractures [20]. Clinical trials that suggested limited therapeutic benefit of HRT and an increased risk of harm (breast cancer, cardiovascular disease) have recently been reassessed and HRT has once again been determined to have a clinical benefit and an acceptable safety profile; despite this, uptake of HRT has declined [20].

The reputational damage to HRT would appear to be such that evidence-based alternative therapeutic options for the prevention of osteoporotic fracture in post-menopausal women are desirable and could have a tangible public health benefit, particularly given the ageing global population [21].

E2, ERα and ERβ are known to play a central role in bone maintenance and turnover - processes that are reliant on a dynamic equilibrium between osteoblast and osteoclast activity. Exogenous signals, e.g. mechanical loading, are sensed by osteocytes. In response, osteoblasts increase their receptor activator of nuclear factor κB ligand (RANKL) expression, which activates pre-osteoclast RANK receptors, thus promoting mature osteoclast formation. Osteoclasts are activated to resorb bone, via the production of hydrogen ions and lysosomal enzymes. At the conclusion of the resorption phase osteoclasts undergo apopotosis and an osteogenic environment dominates once more; crucially osteoblasts express osteoprotegerin (OPG) in such an environment, which serves as a decoy to RANKL, thus limiting the binding of RANKL to its receptor RANK [22]. Broadly, E2 is thought to protect bone mass and architecture by 1) stimulating osteoclast apoptosis and inhibiting apoptosis in osteoblasts and osteocytes; 2) by repressing the production of pro-osteoclastic cytokines (e.g. IL-1, IL-6, TNFα) by osteoblasts; 3) and by increasing the expression of OPG [18,23]. Many of these effects are thought to be mediated via E2 interaction with E2R α [18].

Given the phytoestrogen content of shatavari and evidence of its estrogen-like effects *in vivo* we hypothesised that six weeks shatavari supplementation in postmenopausal women would induce increases in *in vivo* molecular markers of skeletal muscle contractility and protein synthesis. We further hypothesised that sera from shatavari supplemented participants would promote a positive bone turnover phenotype in primary human osteoblasts *in vitro*. Secondarily in this initial six week supplementation study, we measured HGS and knee extensor strength, hypothesising that alterations in muscle contractility might be evident in this timeframe.

## Methods

### Study Participants

This double-blind, placebo-controlled parallel design study was approved by the University of Exeter’s Sport and Health Sciences Research Ethics Committee (190206/B/01). The trial was retrospectively registered at clinicaltrials.gov as NCT05025917 on 30/08/2. All participants gave written informed consent to participate. Twenty-four postmenopausal women (≥ 60 y) were screened and recruited; 20 participants (68.5 ± 6 y, BMI 23.3 ± 3.8) completed the study (study commenced 16/04/2019, participant involvement completed 17/01/2020). In accordance with local ethical approval procedures, participants were free to withdraw without giving a reason; none were withdrawn due to an adverse event. Participants were pre-screened by telephone or email for self-reported exclusion criteria. Exclusion criteria were: BMI > 30; last menstrual period < 12 months previously; use of oestrogenic or progestogenic hormone replacement therapy via oral, transdermal, subcutaneous implant routes in previous five years, diagnosis of osteoporosis or osteopenia, taking any medication for the prevention of osteoporosis, including bisphosphonates and non-bisphosphonates (e.g raloxifene, denosumab, teriparatide, calcitriol). Calcium and vitamin D supplementation were permitted, but no participants reported taking such supplements.

### Study Design and Supplementation Protocol

Participants attended a screening and consent visit at the University of Exeter’s Department of Sport and Health Sciences during which eligibility was again confirmed and informed consent was obtained. Body weight and height were measured. Participants were familiarised with tests of HGS and leg strength, completing the full protocol described in subsequent sections. Participants returned for visit two having fasted overnight and abstained from caffeine consumption in the preceding 24 hours. They completed tests of HGS and leg strength. A blood sample was obtained via venepuncture and a *vastus lateralis (VL)* skeletal muscle biopsy was performed. A researcher who was not involved in data collection or analysis randomised participants to receive either placebo or shatavari supplementation for six weeks. Participants consumed two capsules per day containing either placebo (magnesium stearate, 500 mg/capsule) or shatavari (500 mg powder/capsule equivalent to 13,250 mg fresh weight shatavari root). The capsules were visually identical and did not have an odour. Compliance was assessed by providing an excess of capsules and counting the number that remained at the end of the study period. No participant missed more than 3/42 doses over the six week supplementation period, with 11/20 participants returning the correct number of capsules. Visit three took place six weeks after visit two; the study protocol was repeated as before.

### Measurement of Handgrip Strength

HGS was measured using a handgrip dynamometer according to the Southampton Protocol [24]. Participants sat in a standard chair rested their forearms on the armrest. The wrist was positioned just over the end of the armrest, in a neutral position with the thumb facing upwards. Each hand was tested three times and the tested hand was alternated. The best score for either hand was used for data analysis. One participant reported an injury to their dominant limb during the supplementation period. Their best efforts with their non-dominant limb were included in the data analysis.

### Lower Limb Dynamometry

Assessment of lower limb knee extensor muscle strength was performed using an isokinetic dynamometer (Biodex System 3; Biodex Medical Systems, NY, USA). At the familiarisation visit, participant, chair and dynamometer positioning were personalised, such that the participants sat in the chair with the ankle of their dominant leg fixed to the knee extensor attachment, knee joint in-line with the dynamometer pivot point. Leg dominance was established by asking participants which leg they would preferentially use to kick a ball. Participants’ lower backs were in contact with the chair backrest, which was positioned such that the posterior aspect of their knee had approximately 2 cm clearance from the edge of the seat. Participants were strapped into the chair and instructed to complete all movements with their arms folded across their chests, to isolate the knee extensor effort. All isokinetic movements took place over a range of motion (ROM) of 80 ° from full flexion + 1 ° at the knee, at 60 °·s^-1^ for both the concentric and eccentric phases. A warmup consisting of five sets of five concentric repetitions (1 min rest between sets) was conducted, with participants instructed to complete this at 50 % effort. Following the familiarisation visit, a limit of 60 % isometric maximum voluntary contraction (MVC) was placed on these sets, such the dynamometer would stop a repetition if too much effort was produced. Participants then completed three sets of three maximal isokinetic (IK) repetitions (concentric and eccentric effort; 1 min rest between sets). Following this, the knee was fixed at 90 ° of flexion and three MVCs were performed, separated by 1 min of rest. Force data were recorded and analysed in Spike2 ver.6 software (CED, Cambridge, UK). The best MVC, concentric IK and eccentric IK values attained were used for analysis.

### Blood Sampling and Analysis of Bone Turnover Markers

Antecubital venous blood samples were collected into serum separator (SST) and lithium heparin (LH) containing vacutainer tubes. LH tubes were centrifuged at 4500 rpm for 15 minutes at 4 °C; plasma samples were aliquoted into microcentrifuge tubes. SST tubes were maintained at room temperature for 30 min prior to centrifugation at 4500 rpm for 15 min at 4 °C; serum was aliquoted into microcentrifuge tubes. Plasma and serum samples were stored at – 80 °C until analysis. Carboxy-terminal cross-linking telopeptide of type I collagen (β-CTX) and procollagen type I N-terminal propeptide (P1NP) were quantified using electrochemiluminescence immunoassay on a Cobas e601 analyzer (Roche Diagnostics, Germany), performed at the Bioanalytical Facility, University of East Anglia. The interassay coefficient of variation (CV) for β-CTX was <3 % between 0.01 and 6.0 µg/L, with a sensitivity of 0.01 µng/L. Inter-assay CV for P1NP was <3 % between 5-1200 µg/L with a sensitivity of 5 µg/L.

### Vastus Lateralis Biopsy

*VL* muscle biopsy samples were taken from the exercised (dominant) leg using the suction-modified percutaneous Bergstrom needle technique [25]. The leg from which the biopsy was taken was sterilised with povidone-iodine 10 % w/w cutaneous solution (Ecolab, Northwich, UK) and anaesthetised by infiltration of the skin and subcutaneous adipose tissue with 2 % lidocaine (B Braun, Sheffield, UK). An incision of approximately 0.8 cm was made, and skeletal muscle (∼150 mg) collected. The incision was closed with adhesive butterfly stitches and covered with a waterproof dressing. Skeletal muscle was dissected free of any obvious blood and adipose tissue and immediately snap frozen in liquid nitrogen. Muscle samples were stored at – 80 °C until analysis.

### Immunoblotting

Detailed immunoblotting methods are provided in the Supplementary Material (Supplementary Methods, Supplementary Table 1). Quantifications of protein phosphorylation were normalised to total protein signal. To ensure the veracity of the phosphorylation signals obtained, the same blots were not stripped and re-probed for total and phosphorylated forms of a given protein, rather the electrophoresis and blotting was repeated. Samples were quantified in duplicate. Blot images were never adjusted to manipulate brightness/contrast, except for Coomassie stained blots, where minor adjustments have been made and applied across all lanes.

### Primary Human Osteoblast Cell Culture

Six femoral heads were obtained via the Royal Devon and Exeter Tissue Bank (ethical approval RDETB-REC no 6/SC/0162) which is managed within the NIHR Exeter Clinical Research Facility. Femoral head donors were postmenopausal women (73 ± 9.9 y BMI 29.6 ± 8.6) undergoing hip replacement. Exclusion criteria were: last menstrual period < 12 months previously; use of estrogenic or progestogenic hormone replacement therapy via oral, transdermal, subcutaneous implant routes in previous five years, diagnosis of osteoporosis or osteopenia, taking any medication for the prevention of osteoporosis, including bisphosphonates and non-bisphosphonates (e.g raloxifene, denosumab, teriparatide, calcitriol). Calcium and vitamin D supplementation were permitted. Bone chips were removed from the subchondral region using a Friedman Rongeur and were washed five times in PBS containing 100U/mL penicillin streptomycin. Bone chips were removed to 25 cm^2^ vented flasks containing osteoblast differentiation media: Dulbecco’s Modified Eagle Medium (DMEM) – High Glucose (D6546, Merck, Gillingham, UK) 10 % Fetal Bovine Serum (FBS), qualified E.U. –approved, South America origin (10270106, GIBCO, Waltham, Massachusetts, USA), 100 Units/mL Penicillin/streptomycin (15070-063, GIBCO), 2mM L-Glutamine, G7513 (Merck), 1% Minimum Essential Medium (MEM) Non-essential amino acids, (11140050, GIBCO), β-glycerophosphate disodium salt hydrate (2mM), (G9422, Merck), L-Ascorbic acid (50 µg/ml), (A4403, Merck), Dexamethasone (10 nM), (D4902, Merck). Bone chips were cultured in a humidified incubator (37 °C, 5 % CO_2_). Media were changed after five days and every three days thereafter, with bone chips being removed when osteoblast coverage reached approximately 30 %.

### Assessment of Osteoblast Differentiation

Osteoblasts were seeded at 6×10^3^ cells per well in a 24-well plate and fed with osteoblast differentiation media containing Primary human osteoblasts were stimulated for 14 days with sera (10 % v/v in osteoblast differentiation medium) pooled from eight participants in each supplementation group (baseline and six week timepoints). A 10 % FBS condition was included in each experiment as a previously established control to demonstrate effective differentiation [26]. Osteoblasts were fed by 1:2 demi depletion on days 3, 6, 9 and 12; the experimental endpoint was day 14. All experiments were conducted in four to six biological replicates, with triplicate technical replicates (wells) stimulated with each serum condition for each biologically independent culture. Supernatants were retained for the quantification of protein secretion.

### Osteoblast Cytokine Secretion

Macrophage colony stimulating factor (M-CSF), osteoprotegerin (OPG), interleukin 6 (IL-6) and interleukin 1 beta (IL-1β) were quantified using enzyme-linked immunosorbent assays according to the manufacturer’s instructions (see Supplementary Table 2).

### Osteoblast Bone Nodule Formation

Osteoblast bone nodule formation was quantified via alizarin red staining at day 14 of differentiation. Following removal of the supernatants, osteoblasts were washed three times in phosphate buffered saline (PBS) and were stained with alizarin red staining solution (0.5 % w/v alizarin red (Merck, UK) in 1 % ammonia solution, pH 4.5) for 10 min at room temperature. Osteoblasts were again washed three times in PBS and were then incubated in 10 % w/v cetyl pyridinium chloride (C0732, Merck, UK; warmed to 37 °C) for 10 min at room temperature. This supernatant was further diluted 1:10 in 10 % w/v cetyl pyridinium chloride (warmed to 37°C) and the absorbance of 100 μL of each solution was quantified in duplicate at 550nm on a microplate reader (POLARstar® Omega, BMG LABTECH, Aylesbury, UK).

### Osteoblast Metabolism

Osteoblast metabolism was quantified at day 14 of serum stimulation via an alkaline phosphatase activity assay. Osteoblasts were lysed in RIPA buffer and lysates were diluted 1:4 with 1mM MgCl_2_. Seven alkaline phosphatase standards (3 units/mL top standard, serially diluted 1:2 in 1:4 RIPA:1 mM MgCl_2_) were prepared from a stock solution (Human placenta alkaline phosphatase, P3895, Merck). 10μL sample or standard was added to a 96 well plate in duplicate and 100μL alkaline phosphatase substrate (P7998, Merck) added per well and incubated at 37°C for 15 min. The reaction was stopped by the addition of 0.1N NaOH and the absorbance of each well was measured at 405 nm. ALP concentrations were calculated from the standard curve and corrected for the dilution in MgCl_2_. The protein concentration of the lysates was quantified via BCA assay and alkaline phosphatase activity expressed per unit protein.

### Osteoblast Proliferation

Osteoblast proliferation was quantified via a colourimetric tetrazolium salt-based cell proliferation assay (CellTiter 96® Aqueous One Solution; Promega, Madison, WI, USA). Ostoblast proliferation was assessed at two day intervals, for six days. Osteoblast differentiation medium was replaced with 100 μL serum-free differentiation medium to prevent FBS interference with absorbance readings [27]. 20 μL MTS reagent was added to each well the plate was incubated at 37 °C, 5 % CO2 for 3 h. Each condition was assayed in triplicate. The absorbance of each well was quantified at 450 nm using an absorbance microplate reader.

### Sample Size Calculation and Statistical Analyses

Previous work has demonstrated a 22-fold increase in baseline vastus lateralis myogenin gene expression in post-menopausal woman taking HRT [28]. Myogenin is a molecular marker of terminal myogenic differentiation and thus a marker of skeletal muscle maintenance. However, the magnitude of such gene expression changes is not always reflected in protein expression and the magnitude of such an effect with shatavari supplementation might be expected to be considerably less than that seen with HRT treatment. Thus, we calculated based on a standard deviation of 40% from the mean (unpublished data, calculated SD from n = 9 myogenin western blot quantifications from elderly men) that we would be able to detect a 1.6-fold change in myogenin expression with 11 participants per group. We will therefore recruited 12 participants per group to allow for dropouts. Given the evidence from the literature of an effect of HRT on myogenin expression, and the lack of evidence regarding the effects of shatavari in skeletal muscle, this measure was originally designated as our primary outcome measure. However, the shutdown of our lab facility due to the Covid-19 pandemic lead to the loss of a large number of consumables, including the degradation of many of our immunoblotting antibody stocks that required extensive subsequent troubleshooting. Despite myogenin expression being the designated primary outcome measure, we were forced to use our limited muscle sample stocks to quantify outcomes that required less extensive troubleshooting.

Data are largely quoted mean differences (pre-supplementation to post-supplementation change) for both placebo and shatavari conditions, and have been analysed by unpaired t-test comparison of difference scores and are presented as mean ± standard deviation. Where there exist non-normal distributions, unequal variances or ‘outlier’ data points data were analysed by Mann Whitney U test and presented as median ± interquartile range of difference scores. Data were analysed using GraphPad Prism v5.03 (GraphPad Software, CA, USA). Shapiro-Wilk tests were used to evaluate the normality of data sets and the Levene test was employed to assess the equality of variances within data groups. Outlier data have not been excluded. A p-value of < 0.05 was considered statistically significant.

## Results

### Participant Baseline Strength

Participant baseline strength characteristics are outlined in Supplementary Table 1. There were no significant baseline strength differences between the supplementation groups.

### Shatavari Supplementation Improves Handgrip, but not Knee Extensor Strength in Postmenopausal Women

Six weeks of shatavari supplementation improved HGS in this cohort (placebo -0.4 ± 1.3 kg, shatavari +0.7 ± 1.1 kg; p = 0.04, Mann Whitney U, Figure 1A). Knee extensor isometric (placebo 0 ± 7.8 N, shatavari -4.8 ± 6 N; p=0.14, t-test), concentric (placebo +4.9 ± 10.2 N, shatavari +0.96 ± 6.8 N; p = 0.33, t-test), and eccentric (placebo +2.9 ± 12.1 N, shatavari +5.1 ± 10.7 N; p = 0.67, t-test) strength were not altered by supplement condition (Figure 1 B-C).

**Figure 1.**
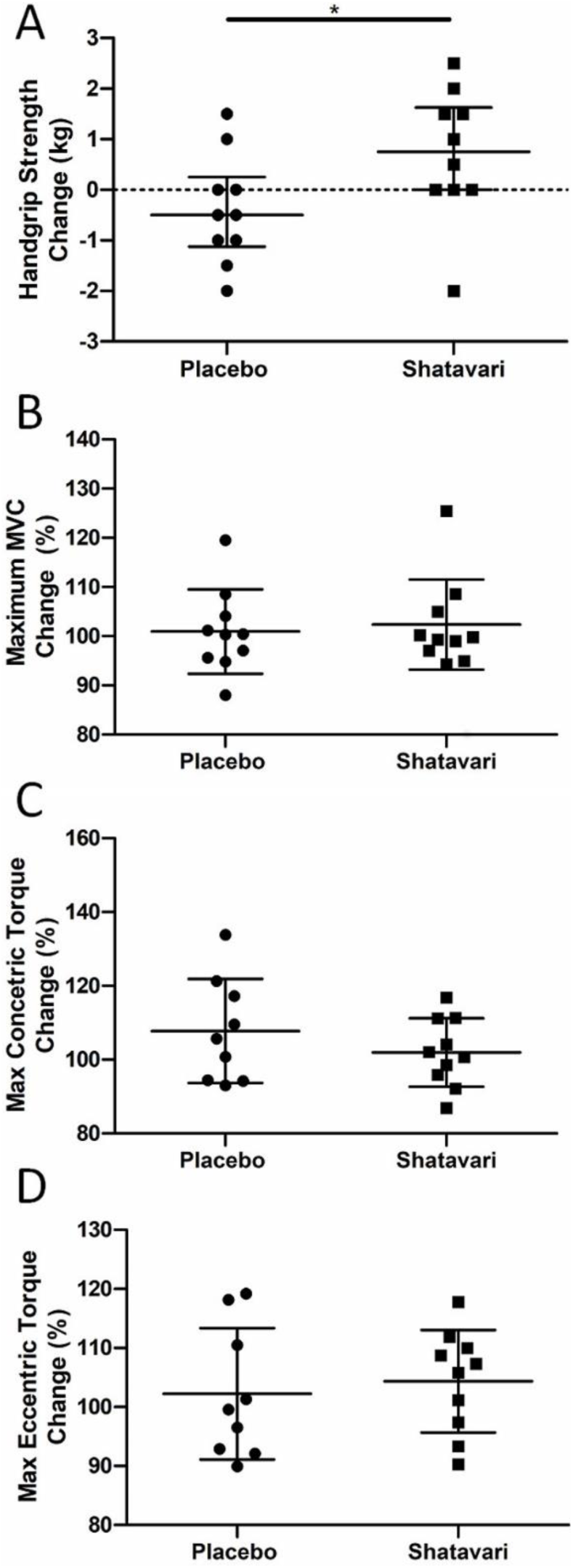
Shatavari Supplementation Improves Handgrip, but not Knee Extensor Strength in Postmenopausal Women. Postmenopausal women supplemented with placebo (N = 10) or shatavari (N = 10) for six weeks. Handgrip strength (A), isometric (B), concentric (C) and eccentric (D) knee extensor strength were measured at baseline and at six weeks. Data are presented as difference scores with the median ± interquartile range (A, outlier present) or mean ± standard deviation (B-D) of these differences. * p < 0.05.

### Shatavari Supplementation Increases Vastus Lateralis Myosin Regulatory Light Chain Phosphorylation in Postmenopausal Women

Myosin regulatory light chain phosphorylation (pMLC) was quantified via immunoblotting and was increased in VL following six weeks of shatavari supplementation (placebo -0.08 ± 0.5 arbitrary units (a.u). shatavari +0.3 ± 1 a.u.; p=0.03 Mann Whitney U) (Figure 2). An additional panel demonstrating the spread of data with the lowest datapoint in the placebo dataset removed can be found in the Supplemental Material (Supplemental Figure 1).

**Figure 2.**
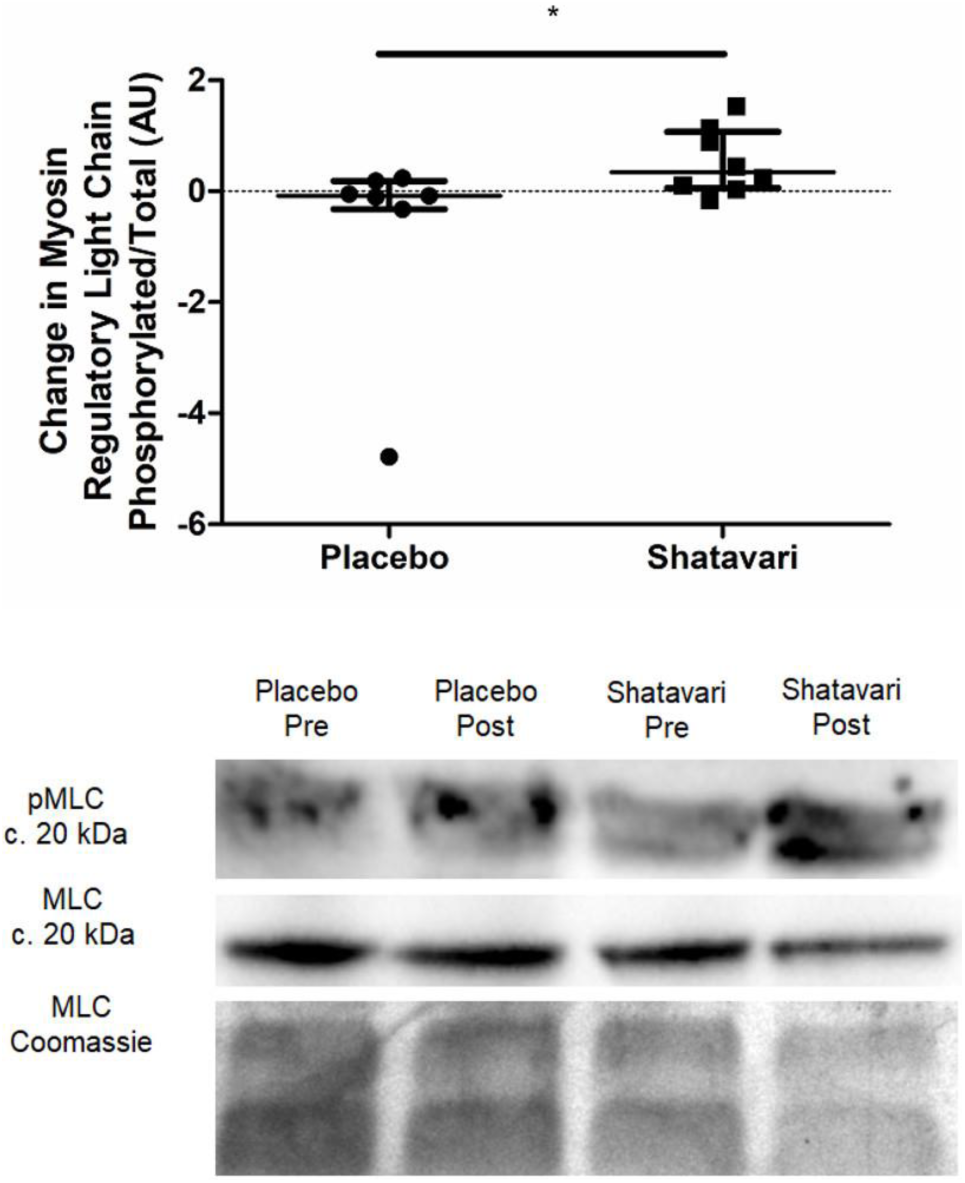
Shatavari Supplementation Increases *Vastus Lateralis* Myosin Regulatory Light Chain Phosphorylation in Postmenopausal Women. Postmenopausal women supplemented with placebo (N = 10) or shatavari (N = 10) for six weeks. Total myosin regulatory light chain (MLC) and myosin regulatory light chain phosphorylation (pMLC) were quantified via immunoblotting at baseline and at 6 weeks. Samples were assayed in duplicate. Data are presented as difference scores with the median ± interquartile range of these differences. Missing data are accounted for by failed muscle biopsy/biopsy sample of insufficient quantity.

### Shatavari Supplementation has an Equivocal Effect on Vastus Lateralis Protein Synthetic Pathways

The phosphorylation status of protein synthetic pathway effectors was quantified via immunoblotting at baseline and following six weeks of shatavari supplementation. Shatavari increased phosphorylation of Akt^ser473^ (placebo -0.6 ± 0.6 a.u. shatavari +0.2 ± 1.3 a.u; p=0.03 Mann Whitney U). Phosphorylation of p70S6k^Thr389^, 4EBP1^Thr37/46^,S6^Ser240/244^ did not differ between conditions (Figure 3).

**Figure 3.**
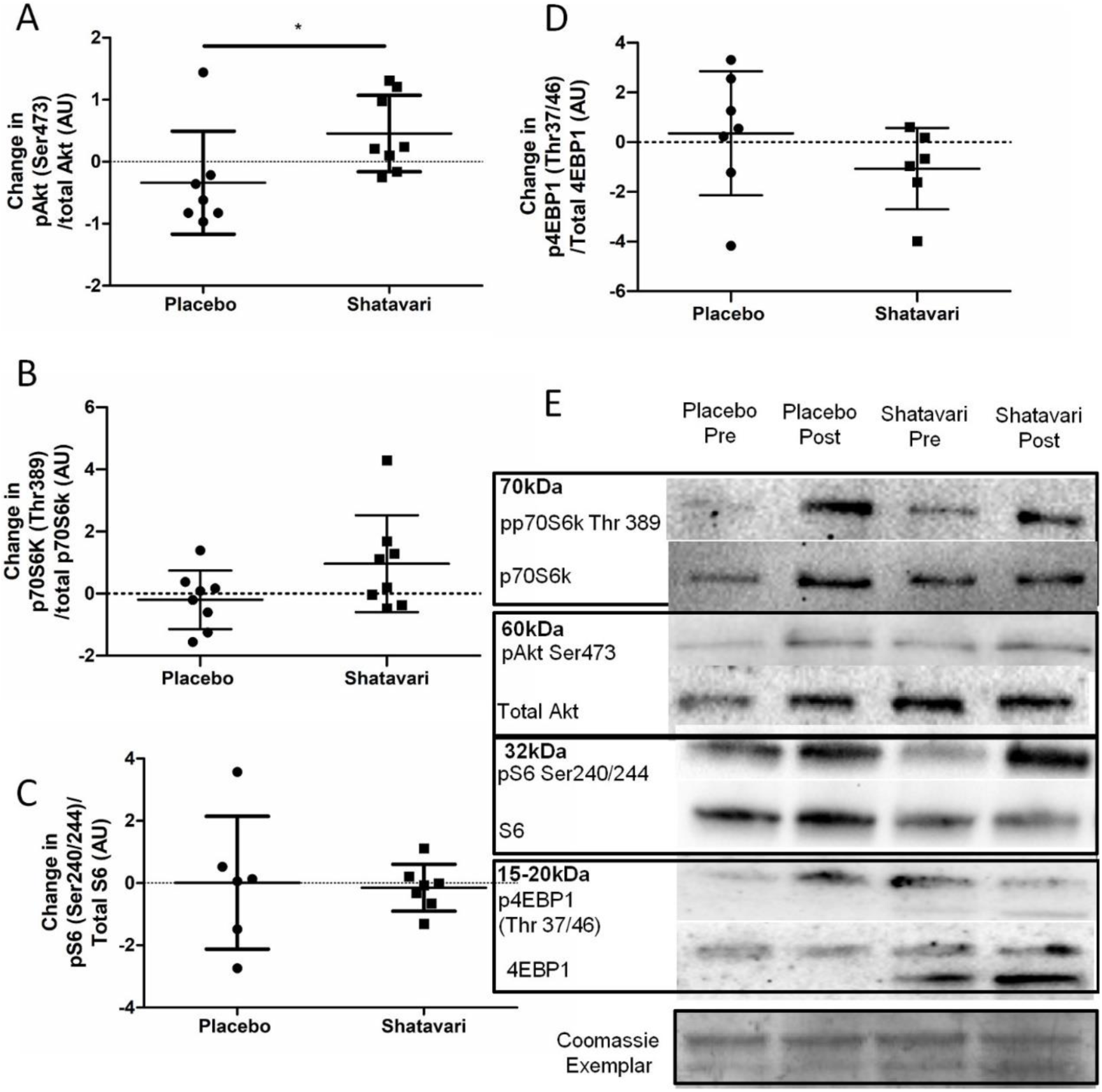
Shatavari Supplementation has an Equivocal Effect on *Vastus Lateralis* Protein Synthetic Pathways. Postmenopausal women supplemented with placebo (N = 10) or shatavari (N = 10) for six weeks. Phosphorylation of Akt^ser473^ p70S6k^Thr389^, 4EBP1^Thr37/46^,S6^Ser240/244^ were quantified via immunoblotting at baseline and at six weeks. Samples were assayed in duplicate. Data are presented as difference scores with the median ± interquartile range (A, C, D) or mean ± standard deviation (B) of these differences. * p < 0.05. Missing data are accounted for by failed muscle biopsy/biopsy sample of insufficient quantity.

### Shatavari Supplementation Does Not Alter In Vivo or In Vitro Markers of Bone Turnover

Plasma concentrations of bone turnover markers P1NP and β-CTX were quantified at baseline and following six weeks of shatavari supplementation. Neither P1NP (placebo -0.4 ± 8.7 μg/L, shatavari + 1.3 ± 6.5 μg/L; p=0.16, t-test), nor β-CTX (placebo +0.01 ± 0.11 μg/L, shatavari -0.08 ± 0.22 μg/L; p=0.27, Mann Whitney U), were altered by supplementation condition (Figure 4 A, B). Primary human osteoblasts (N=6) were stimulated for 14 days with sera (10% in osteoblast differentiation medium) from participants in both supplementation groups (baseline and six week timepoints). Supernatant concentrations of osteoblast-produced endocrine/paracrine cytokines were quantified by ELISA. Concentrations of OPG (placebo +0.24 ± 2.61 ng/mL, shatavari + 0.01 ± 2.94 ng/mL; p=0.89, t-test), M-CSF (placebo +55.7 ± 100.5 ng/mL, shatavari -4.7 ± 67.0 μg/L; p=0.25, t-test), IL-6 (placebo +58.8 ± 284.9 pg/mL, shatavari +189.6 ± 344.5 pg/mL; p=0.39, Mann

**Figure 4.**
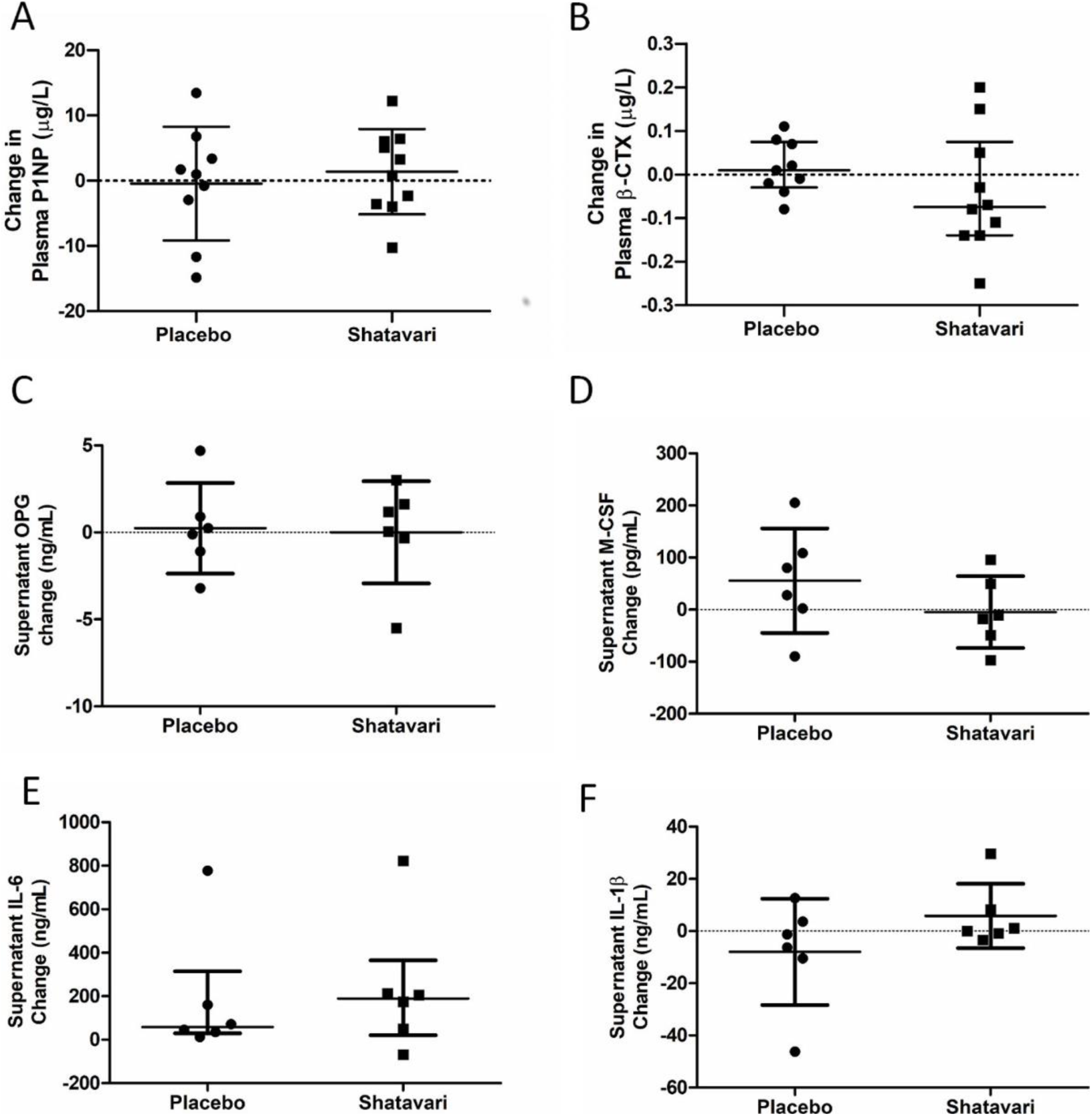
Shatavari Supplementation Does Not Alter Plasma Markers of Bone Turnover or Osteoblast Cyokine Secretion. Postmenopausal women supplemented with placebo (N = 10) or shatavari (N = 10) for six weeks. Plasma concentrations of bone turnover markers Procollagen 1 Intact N-Terminal Propeptide (P1NP, A) and beta-C-terminal telopeptide (β-CTX, B) were quantified at baseline and following six weeks of supplementation. Primary human osteoblasts were stimulated for 14 days with sera (10 % in osteoblast differentiation medium) pooled from eight participants in each supplementation group (baseline and six week timepoints). Supernatant concentrations of osteoblast-produced endocrine/paracrine cytokines were quantified by ELISA. C) osteoprotegerin (OPG) D) macrophage colony stimulating factor (M-CSF) E) interleukin 6 (IL-6); F) interleukin 1 beta (IL-1β). Data are presented as difference scores with the median ± interquartile range (B, D) or mean ± standard deviation (A, C, E, F) of these differences.

Whitney U), IL-1β (placebo -8.0 ± 20.4 ng/mL, shatavari +5.8 ± 12.3 ng/mL; p=0.19, t-test), were not altered by supplementation condition (Figure 4 C-F). TNFα could not be detected.

Alkaline phosphatase activity was quantified as an indicator of osteoblast biosynthetic activity, with no difference between supplementation conditions being observed (placebo -0.01 ± 0.31 units activity/mg protein, shatavari +0.04 ± 1.80 μg/L; p=0.22, Mann Whitney U) (Figure 5 A). Oestrogens are known to increase osteoblast proliferation [29]. MTS assay of proliferating osteoblasts showed that there was no significant difference in mean MTS absorbance between supplementation conditions, (F(3, 24) = 1.027, p = 0.42) (Figure 5 B). Alizarin red staining detected no difference in osteoblast-produced calcium deposits between placebo and shatavari sera conditions (absorbance change pre to post: placebo +0.07 ± 0.40, shatavari +0.07 ± 0.58; p=0.22, Mann Whitney U) (Figure 5 C). No difference between supplementation conditions was observed for expression of protein markers of osteoblast differentiation (Figure 5 D-F).

**Figure 5.**
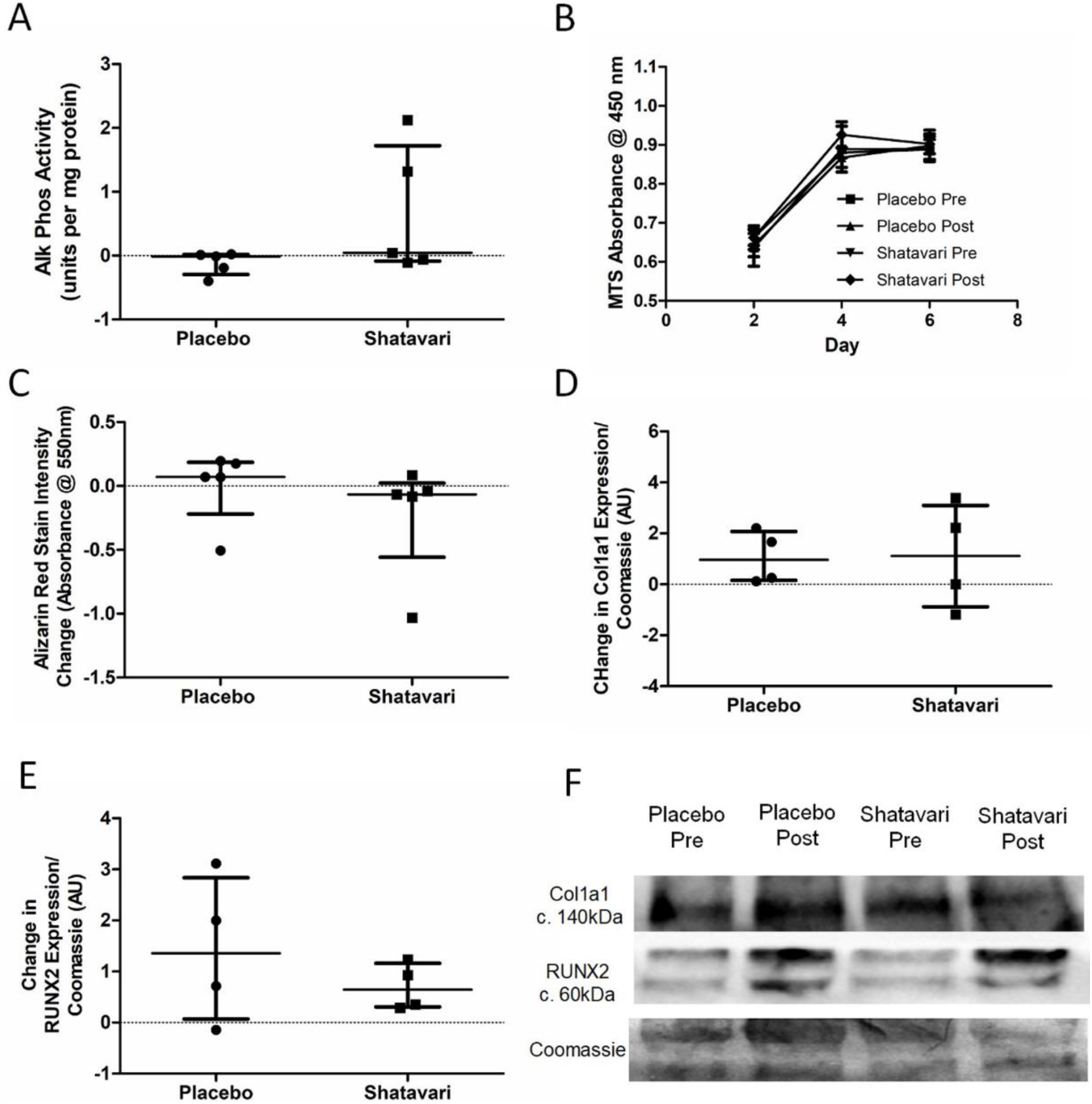
Shatavari Supplemented Sera Do Not Alter Osteoblast Metabolic Activity or Differentiation. Primary human osteoblasts were stimulated with sera (10 % v/v in osteoblast differentiation medium, 14 days for all except B – 6 days stimulation) pooled from eight participants in each supplementation group (shatavari/placebo, baseline and six week timepoints). A) Alkaline phosphatase activity B) Cellular proliferation rate C) Bone nodule formation (Alizarin Red stain intensity) and D-F) differentiation markers (Col1a1, RUNx2) were quantified. Data are presented as difference scores with the median ± interquartile range (A-D) or mean ± standard deviation (D,E) of these differences.

## Discussion

This is the first study to assess the effect of shatavari on skeletal muscle function in a cohort of older adults and the first offer insights into the molecular changes that may underpin its functional effects. To our knowledge, this is also the first study to assess the effect of shatavari supplementation on markers of bone turnover. Six weeks of shatavari supplementation improved HGS in this cohort of postmenopausal women. Although no improvement in knee extensor strength was found, we demonstrated an increase in pMLC within the *VL*. We also demonstrated an increase in Akt^Ser473^ phosphorylation, although no alteration in phosphorylation status of other major effectors of muscle protein synthesis (p70S6k^Thr389^, 4EBP1^Thr37/46^, S6^Ser240/244^) was observed. We did not find any effect of shatavari supplemented sera on markers of primary human osteoblast function.

An increase in HGS and pMLC over a period of six weeks is congruent with what is known about the effect of E2 and pMLC on muscle strength. Such phosphorylation modulates striated muscle contraction by promoting movement of the myosin head towards the thin filament, thus increasing the probability of myosin interaction with actin [30–32]. Age -dependent reductions in pMLC have been documented and appear to particularly affect women [33,34]. Our observation that shatavari increased pMLC and muscle strength over six weeks is consistent with the time course of such changes documented elsewhere; Lai et al. showed that E2 administration over 4-6 rescued extensor digitorum longus and soleus pMLC and force generation in bilaterally ovariectomized mice [35]. Further, deletion of E2R α reduces muscle contractility and force production in female mice [36]. More work is required to definitively establish the link between shatavari-induced pMLC changes and human skeletal muscle contractility e.g. via *ex vivo* assessment of muscle fibre contractility and using *in vivo* techniques such as peripheral nerve stimulation (PNS) and transcranial magnetic stimulation (TMS) to assess central drive, excitability and muscle contractility.

Perhaps surprisingly in the context of the shatavari-induced changes in HGS, knee extensor strength was not increased despite an increase in *VL* pMLC. Our subjective experience during this study was that the learning effect of our familiarisation session for this older adult cohort was not as effective as the same familiarisation protocol that we use regularly in younger cohorts. Indeed, there may be greater variability in reaching maximum voluntary activation in older age and our ‘best of three’ approach to such tests [37] may need to be modified in such cohorts to take the best value from a greater number of attempts [38]. Testing of HGS is an inherently more ‘accessible’ movement for those unfamiliar with resistance exercise and our *a priori* decision to use the best value from all six trials (three dominant, three non-dominant) in accordance with the ‘Southampton Protocol’ for such testing may have mitigated this variability [24]. Indeed, the coefficient of variation (CV) for the best HGS scores achieved at visits 2 and 3 for the placebo group was 2.9 ± 2.3 %; the CV for the best MVC value was 4.1 ± 4 %, although this did not reach statistical significance (p = 0.46, t-test). Notably the CVs for concentric (8.2 ± 6.5 %) and eccentric (6.6 ± 4.1 %) isokinetic measures of muscle strength were significantly larger than that for HGS (p = 0.03, t-test for both). The lack of a forearm skeletal biopsy in the context of our HGS observations is unfortunate, however there the *VL* remains the most accessible skeletal muscle for biopsy outside of a secondary/tertiary medical care facility [39].

Intriguingly, we also observed an increase in Akt^Ser473^ phosphorylation, but no alteration in phosphorylation status of other major effectors of muscle protein synthesis signalling. Interestingly in the context of this increase in Akt^Ser473^ phosphorylation, it has been suggested that one way in which pMLC may be increased is via the PI3K/MAPK/Akt/MLCK pathway, [35]. However, no definitive activation of the proteins downstream of mTOR was observed. We note however that our analysis of p70S6k^Thr389^ showed an increase in its expression with shatavari supplementation with p = 0.09. We do not consider non-significant results such as this worthy of excessive weighting in a discussion of results or in the drawing of conclusions. However, we highlight this result in the context of the relatively low number of viable biopsy samples that were available for analysis. We powered our analyses (using standard deviations derived from unpublished immunoblotting data from older adults) to be able to detect a 1.6-fold change in protein expression with 10 participants per group; we recruited 12 participant per group to account for dropouts (two dropouts occurred per group) and biopsy failures. However, even with a ‘second pass’ following a failed biopsy, we had a muscle tissue extraction failure rate of 7.5% in this population, thus these analyses did not have the statistical power that we had anticipated. Work published since the conception of our study has shown that c. 40 % of healthy older female participants are at risk of a failed muscle biopsy. Wilson et al. highlight low muscle mass as the principal contributor to this failure [40]. Our samples also often contained a high proportion of subcutaneous or intramuscular adipose tissue, which accumulates in skeletal muscle with advancing age [41]. Despite these caveats, we have demonstrated novel functional effects of shatavari supplementation in skeletal muscle, along with some important mechanistic insights. We consider that the potential for shatavari supplementation to enhance muscle protein synthesis should be further explored in longer-term resistance training studies, given our evidence of Akt^Ser473^ phosphorylation following shatavari supplementation, coupled with the observations of Anders et al. that shatavari enhanced strength gains in young men following eight weeks of bench press training [17]. Indeed, the balance of current evidence suggests that E2 increases the anabolic response to exercise over the longer term (for an excellent review see [42]. It remains to be seen what sex differences exist in the effect of shatavari on skeletal muscle function. The work of Anders et al. suggests that its effects are not limited to the female sex, despite the primacy of testosterone and its derivatives as endocrine determinants of male skeletal muscle function [43]. It should be borne in mind that shatavari also contains steroidal saponins, racemosides, racemosol, racemofuran and asparagamine A; little is known about their effects on skeletal muscle.

Indeed, the chemical complexity of shatavari and its metabolism *in vivo* is another area that warrants extensive further research. Determining the constituents of shatavari that evoke physiological responses is not a simple task. The bioavailability of phytoestrogen-containing supplements is variable and poorly understood. The intestinal flora produce both aglycone primary metabolites and secondary metabolites that potentially have a greater affinity for E2R than their parent compounds [44]. Indeed, this is why primary human osteoblasts in this study were treated with sera from women who had supplemented with placebo or shatavari for six weeks, with their final dose taken one hour prior to venepuncture (i.e. an acute-on-chronic serum supplementation condition). *In vitro* studies of bioactive supplements almost universally apply extracts of the supplements directly to cell cultures. For the reasons outlined above, we consider this approach to be inappropriate and lacking in physiological relevance.

The decrease in E2 production that potentiates sarcopenia in women after the menopause is also associated with a decline in bone mineral density. Given this observation, we also examined systemic and *in vitro* markers of bone turnover. Here, we utilised a previously established model for assessing metabolic activity and bone nodule formation in human primary osteoblasts [26]. We did not find any effect of shatavari supplemented sera on markers of primary human osteoblast function, neither did we observe any changes in the osteoblast secretion of cytokines that are known to alter osteoclast activity. In this six week supplementation study, these *in vitro* assays of bone turnover activity were the bone outcomes of primary interest and considered the most likely to be altered by supplement condition. However, we also quantified *in vivo* markers of bone turnover (P1NP and β-CTX), with no effect of supplementation being observed. Given the *in vitro* results that we describe, this is most likely due to a genuine lack of effect on shatavari on bone turnover. However, despite best efforts *in vitro* models cannot perfectly reproduce human physiology and it remains possible that shatavari supplementation could have an effect on bone turnover in the longer term. Transdermal E2 treatments do induce changes in such markers over a similar timeframe. However, within-subject variability for these measures can be high (least significant change P1NP 21%, β-CTX 132%) [45] and the potency of any effect of shatavari on such measures might be expected to be less than pharmacological doses of E2.

## Conclusions

Here, we examined the potential benefits of shatavari supplementation for the musculoskeletal system after the menopause and this initial study has signposted several novel effects and promising avenues for further research in this area. Discussion of the limitations of our approach and conclusions has been integrated into the main discussion. Specifically, we found that shatavari appears to improve muscle function, potentially mediated by improvements in myosin contractility and warrants further investigation of its utility in conserving musculoskeletal function in older adults. Further, the constituents of shatavari that mediate these effects – estrogenic or otherwise – require identification.

## Supporting information

Conflict of Interest Disclosure

CONSORT checklist

## Data Availability

The datasets used and/or analysed during the current study are available from the corresponding author on reasonable request.

## Declarations

### Ethics approval and consent to participate

This study was approved by the University of Exeter’s Sport and Health Sciences Research Ethics Committee (190206/B/01). All participants gave written informed consent to participate.

## Authors’ contributions

MOL, JB conceived and planned the experiments. MOL, JB, VS, MC, SJ, JT, JD carried out the experiments. MOL, JB contributed to the interpretation of the results. MOL took the lead in writing the manuscript. All authors provided critical feedback and helped shape the research, analysis and final manuscript. All authors have approved the submitted version. All authors have agreed both to be personally accountable for the author’s own contributions and to ensure that questions related to the accuracy or integrity of any part of the work, even ones in which the author was not personally involved, are appropriately investigated, resolved, and the resolution documented in the literature.

## Acknowledgements

We thank Dr Bea Knight, Ms Lidia Romanczuk and Mr John Charity for their assistance with this research project. We thank Dr Vivien Rolfe, Marion Mackonochie and Simon Mills from Pukka Herbs for useful discussions regarding the Ayurvedic uses of shatavari. This project was supported by the National Institute for Health Research (NIHR) Exeter Clinical Research Facility which is a partnership between the University of Exeter Medical School College of Medicine and Health, and Royal Devon and Exeter NHS Foundation Trust. The views expressed are those of the author (s) and not necessarily those of the NIHR or the Department of Health and Social Care.

