## Supplemental Material for "Shatavari Supplementation in Postmenopausal Women Improves Handgrip Strength and Increases *Vastus Lateralis* Myosin Regulatory Light Chain Phosphorylation But Does Not Alter Markers of Bone Turnover: A Randomised Controlled Trial"

**Supplementary Methods**

**Immunoblotting**

20 mg skeletal muscle was homogenised in screw top microcentrifuge tubes containing 3 steel beads (3.2mm diameter) and 400μL ice-cold radioimmunoprecipitation assay (RIPA) buffer (50 mM Tris HCl, 150 mM NaCl, 1.0 % (v/v) NP-40, 0.5 % (w/v) Sodium Deoxycholate, 1.0 mM EDTA, 0.1 % (w/v) sodium dodecyl sulfate [SDS], 0.01 % (w/v) sodium azide, protease inhibitor cocktail, phosphatase inhibitor cocktail, pH of 7.4). Homogenisation was performed using a Speedmill Plus homogeniser (Analytik Jena AG, Jena, Germany). Lysates were then removed to a different microcentrifuge tube and centrifuged at 14,000 g for 10 min at 4 °C. The supernatant was removed to another microcentrifuge tube. The total protein concentration of each cell lysate supernatant was quantified by a bicinchoninic acid assay (Thermo Scientific), according to the manufacturer's instructions. Lysates were stored at - 80 °C prior to use in downstream applications.

Total protein lysates of known concentration were mixed 3:1 with 4x Laemmli sample loading buffer (0.5M Tris-HCl pH 6.8, 20 % (v/v) Glycerol, 10 % (w/v) SDS, 0.1 % (w/v) bromophenol blue, 10 % (v/v) beta-mercaptoethanol [βME]) and water to generate polyacrylamide gel loading stocks of known concentration. Samples were boiled at 100°C for 5 min. Pre-cast polyacrylamide gels (Mini-PROTEAN® TGX™ Precast Gels, Bio-Rad) were loaded with an appropriate volume of loading stock (see Supplementary Table 1 for details of loading mass, gel percentages etc). Samples from both supplementation conditions were run side by side on each gel.

Proteins were transferred onto polyvinylidene difluoride (PVDF) membrane (Amersham Hybond-P, GE Healthcare, Buckinghamshire, UK) in an electroblotting buffer (0.025M Tris base, 0.192M glycine, 20 % (v/v) methanol at pH 8.4) using the 'Standard SD' program of the Trans-Blot Turbo semi-dry transfer system (Bio-Rad, CA, USA). Membranes were blocked for 1 h and agitated overnight with primary antibody diluted in TBS-T at 4 °C (Table). The membranes were washed for 4 x 5 min in 1xTBS-T (50 mM Tris hydrochloride [Tris-HCl], 150 mM sodium chloride [NaCl], 0.1% (v/v) Tween 20 at pH 7.5) and agitated for 1 h at room temperature with horseradish peroxidase-conjugated secondary antibody (Table). TBS-T washes were repeated and the membranes incubated with ECL Prime Western blotting detection reagents (Amersham Biosciences, Amersham, UK) according to the manufacturer's instructions. Bands were visualised on the ChemiDoc MP imaging system (Bio-Rad). Blots were washed briefly in TBS-T and stained with 0.1% (w/v) coomassie blue solution (Methanol (50% [v/v], glacial acetic acid 10% [v/v] H2O 40% [v/v]) to ensure even protein loading. An open-source, public domain software package (Image J v1.47) was used for band quantification.

**Supplementary Table 1: Immunoblotting Antibody Details**

| Target Protein | SDS Page Gel % | Protein  Load (µg) | Blocking Solution (5% w/v) | Primary Antibody Details (catalogue number, supplier) | Primary Antibody Dilution in TBS-T | | Secondary Antibody Details/ Dilution in TBS-T |
| --- | --- | --- | --- | --- | --- | --- | --- |
| Akt total | 12% | 20 | Milk | Akt (9272, CST) | | 1:3000 | Goat Anti-Rabbit IgG H&L (HRP) (ab205718) IgG antibody (Abcam, Cambridge, UK)  1: 15000 |
| Akt Ser^473^ | 12% | 20 | BSA | Phospho-Akt Ser473 (4060T, CST) | | 1:3000 |  |
| p70S6k | 12% | 20 | Milk | p70S6k (9202, CST) | | 1:5000 |  |
| p70S6k^Thr389^ | 12% | 20 | BSA | Phospho-p70 S6 Kinase Thr389 (9234, CST) | | 1:5000 |  |
| 4EBP1 | 4-20% | 30 | Milk | 4EBP1 (9452, CST) | | 1:2500 |  |
| 4EBP1^Thr37/46^ | 4-20% | 30 | Milk | Phospho-4E-BP1 Thr37/46 (9459, CST) | | 1:2500 |  |
| S6 | 4-20% | 30 | Milk | S6 Ribosomal Protein (5G10, CST) | | 1:2500 |  |
| S6Ser^240/244^ | 4-20% | 30 | Milk | Phospho-S6 Ribosomal Protein Ser 240/244 (5364, CST) | | 1:2500 |  |
| MLC | 12% | 30 | Milk | Anti-Myosin Light Chain 2 antibody (ab79935, Abcam) | | 1:2500 |  |
| MLC^ser20^ | 12% | 30 | Milk | Anti-Myosin light chain (phospho S20) antibody (ab2480, Abcam) | | 1:2500 |  |
| Col1a1 | 4-20% | 20 | Milk | Anti-COL1A1 antibody [N1N2], N-term (GTX112731, Genetex) | | 1:2500 |  |
| RUNX2 | 4-20% | 20 | Milk | RUNX2 (#12556, CST) | | 1:2500 |  |

Sodium dodecyl-sulphate polyacrylamide gel electrophoresis (SDS-PAGE), Tris-buffered saline with 0.1% Tween ® 20 Detergent (TBS-T), Bovine Serum Albumin (BSA), Horseradish peroxidase (HRP), Cell Signalling Technology (CST)

**Enzyme Linked Immunosorbent Assays**

**Supplementary Table 2: ELISA Details**

| Target Protein | Product Name | Catalogue Number | Manufacturer |
| --- | --- | --- | --- |
| M-CSF | Human M-CSF DuoSet ELISA | DY216 | R&D Systems |
| OPG | Human Osteoprotegerin/TNFRSF11B DuoSet ELISA | DY805 | R&D Systems |
| IL-6 | IL-6 Human Uncoated ELISA Kit | 88-7066-22 | 20 |
| IL-1β | IL-1 beta Human Uncoated ELISA Kit | 88-7261-22 | 20 |

Macrophage colony-stimulating factor (M-CSF), osteoprotegerin (OPG), interleukin 6 (IL-6), interleukin 1 beta (IL-1β)

**Supplementary Results**

**Participant baseline outcomes**

Participant baseline strength characteristics are outlined in Supplementary Table 3. There were no significant baseline strength differences between the supplementation groups.

**Supplementary Table 3: Participant Baseline Strength**

| Outcome | Placebo | Shatavari | P value |
| --- | --- | --- | --- |
| Handgrip Strength (kg, mean ± SD) | 23.1 ± 4.2 | 21.7 ± 5.0 | 0.5 (t-test) |
| Maximum Voluntary Contraction (N mean ± SD) | 104.9 ± 31.1 | 109.0 ± 16.4 | 0.72 (t-test) |
| Maximum Isokinetic Concentric Force (N median ± IQR) | 81.1 ± 46.4 | 82.0 ± 21.9 | 0.85 (Mann Whitney U) |
| Maximum isokinetic Eccentric Force (N median ± IQR) | 111.1 ± 27.1 | 106.7 ± 31.6 | 0.97 (Mann Whitney U) |

**Myosin Regulatory Light Chain Phosphorylation**

The spread of myosin regulatory light chain phosphorylation data with the lowest datapoint in the placebo dataset removed is presented in Supplementary Figure 1. These data are included to allow the reader to appreciate the spread of other data. No valid reason for exclusion of the lowest datapoint from analyses was identified.

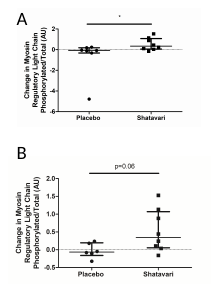

**Supplementary Figure 1. Shatavari Supplementation Increases Vastus Lateralis Myosin Regulatory Light Chain Phosphorylation in Postmenopausal Women. Postmenopausal women supplemented with placebo (N = 10) or shatavari (N = 10) for 6 weeks**. Total myosin regulatory light chain (MLC) and myosin regulatory light chain phosphorylation (pMLC) were quantified via immunoblotting at baseline and at 6 weeks. Samples were assayed in duplicate. Data are presented as difference scores with the median ± interquartile range of these differences. A) Full dataset B) spread of data with the lowest datapoint in the placebo dataset removed.

**Full Versions of Immunoblots presented in the Manuscript**

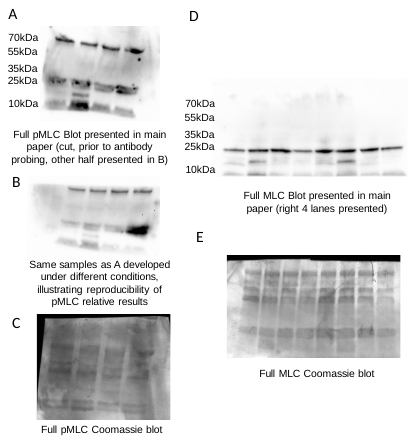

**Supplementary Figure 2. Full Versions of Myosin Light Chain Immunoblots Presented in the Manuscript**

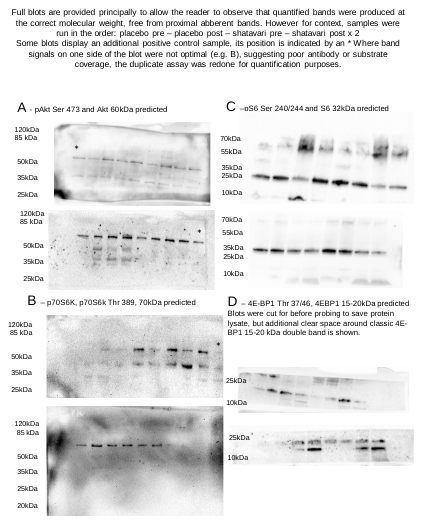
**Supplementary Figure 3. Full Versions of Other Immunoblots Presented in the Manuscript**
